# Novel autoantibodies in patients with systemic sclerosis and gastrointestinal dysfunction provide insight into disease pathogenesis

**DOI:** 10.1101/2025.08.05.25333061

**Authors:** Zsuzsanna H. McMahan, Ami A. Shah, Srinivas N. Puttapaka, Livia Casciola-Rosen, Subhash Kulkarni

## Abstract

**Objective:** Gastrointestinal (GI) dysfunction in systemic sclerosis (SSc) is common and debilitating, yet its underlying mechanisms and related biomarkers are poorly understood. We sought to discover novel autoantibodies in patients with SSc-GI dysfunction and evaluate their clinical relevance.

**Methods:** Sera from 111 SSc patients enrolled in the Gastrointestinal Assessment Protocol (GAP) were screened for novel autoantibodies. Using immunoprecipitation of murine myenteric plexus lysates followed by mass spectrometry, autoantibodies targeting Argonaute RISC Catalytic Component 1/2 (AGO) and Dihydrolipoamide Branched Chain Transacylase E2 (DBT) were identified. Clinical associations were evaluated in two SSc cohorts. Expression of AGO and DBT in the murine enteric nervous system (ENS) was confirmed by immunohistochemistry. ENS-derived extracellular vesicles (EVs) from longitudinal muscle-myenteric plexus tissues were analyzed for AGO cargo using flow cytometry and western blotting.

**Results:** Anti-AGO antibodies occurred in 13.5% and anti-DBT in 4% of GAP patients. Anti-AGO antibodies were associated with severe constipation on the UCLA GIT 2.0 (26% vs. 9%, p=0.036), and anti-AGO2, specifically, associated with severe distention and bloating (16% vs. 4%, p=0.046). Higher AGO1/2 antibody levels associated with severe constipation (p=0.03). Anti-DBT patients exhibited less esophageal emptying at 10 seconds (30% vs. 81%, p=0.034) and less constipation [median 0 (IQR 0–0) vs. 0.75 (0.25–1), p=0.02]. Immunofluorescence studies revealed that anti-AGO antibodies target AGO2-containing gut-derived EVs, whereas anti-DBT antibodies recognize mesoderm-derived enteric neurons and smooth muscle.

**Conclusion:** SSc patient autoantibodies may reveal distinct clinical phenotypes and disease mechanisms that can define biomarkers, disease pathways, and targets for potential therapeutic strategies.

## INTRODUCTION

Gastrointestinal (GI) dysmotility is a common and debilitating consequence of systemic sclerosis (SSc) that impacts function and quality of life in most patients^1^. A significant part of GI motility is regulated by the enteric nervous system (ENS), whose neurons reside within the gut wall^2^. Prior data demonstrate that the SSc autoimmune response targets enteric neurons, and that functional autoantibodies to neurons and smooth muscle play an important role in SSc-GI dysmotility and inform GI risk stratification in ∼10% of SSc patients^3, 4^. Our group previously demonstrated that autoantibodies in sera from patients with SSc GI disease target major ENS components, including mitochondrial antigens and cell membrane proteins^4, 5^. Furthermore, functional antibodies to muscarinic-3 receptors (M3R) target cell surface receptors on neurons and GI smooth muscle which play a role in the development of rapidly progressive GI disease and pseudo-obstruction^3, 6^.

While GI dysmotility is common in SSc, and data suggest that some patients have anti- M3R antibodies contributing to their GI dysfunction, overall disease mechanisms remain poorly understood and existing therapies are therefore not disease-modifying. Furthermore, symptom control does not effectively prevent GI progression, and there are no markers of disease activity. An improved understanding of the key drivers of SSc GI dysmotility, and the mechanisms that contribute to persistent disease activity and progression are critical in identifying high-risk patients and learning how to modify disease course.

Our group previously found that autoantibodies in sera from SSc patients target novel autoantigens expressed by enteric neurons, with a subset showing preferential binding to the newly discovered lineage of mesoderm-derived enteric neurons (MENs)^7^. This is noteworthy because relevant targeted antigens may be more abundant in MENs than in the canonical lineage of neural crest-derived enteric neurons (NENs)^4^. Since this recently described subgroup of ENS neurons is thought to play a critical role in regulating important GI functions, including GI motility, we sought to identify additional autoantigen(s) expressed by enteric neurons - specifically by MENs - that are targeted by autoantibodies in SSc patients with GI dysmotility, and to define the clinical features (GI and extra-intestinal) associated with the identified specificities.

## METHODS

### Patient Cohorts

#### SSc All-Comers Cohort

This consisted of all SSc patients regardless of GI disease status (N=178) recruited consecutively at the Johns Hopkins Scleroderma Center during routine clinical visits from April 2016 to August 2017^5^. The Johns Hopkins Scleroderma Center Research Registry contains demographic and detailed clinical data from patients at their first clinical encounter and every 6 months thereafter^8^. Evidence of patient-reported GI symptoms was determined by the maximum UCLA SCTC GIT 2.0 survey score^9^. Significant GI disease was defined as moderate to severe GI severity and categorized by the UCLA GIT 2.0 score, using cutoffs as previously described^10^.

#### SSc Gastrointestinal Assessment Protocol (GAP) Cohort

This consisted of SSc patients with more severe GI symptoms and whole gut transit testing (N=111) as previously described^11, 12^.

#### Healthy Controls

Serum was obtained from healthy controls (N=38) without SSc or other known autoimmune diseases.

All human subjects protocols were approved by the Johns Hopkins IRB, with informed consent obtained from all participants.

#### Isolation and culture of the longitudinal muscle containing myenteric plexus (LM-MP) from adult murine small intestine (ileum)

LM-MP tissue extracts were prepared as described before^5^. Ileal LM-MP tissues from adult male C57BL/6 wildtype mice and from Wnt1- cre:tdTomato lineage fate mapping mice, which we have previously used to label all neural crest-derivatives in the GI tract with the red fluorescent reporter tdTomato, were isolated as described^4^. LM-MP tissues were fixed for 5 minutes with ice-cold 4% paraformaldehyde within 30 minutes of isolation, after which they were blocked, permeabilized, and immunostained with appropriate antibodies. For interrogating AGO2 immunoreactivity, LM-MP tissues from C57BL/6 wildtype mice were immunostained with anti-AGO2 antibody (Cell Signaling, Cat. No. 2897) and with ANNA1 serum containing autoantibodies against the neuronal Hu proteins (to identify myenteric ganglia, ANNA1 serum was used diluted 1:1000). For interrogating anti-DBT immunoreactivity, LM-MP from Wnt1-cre:tdTomato lineage fate mapping mice were immunostained with anti-DBT antibody (Gentex, Cat. No. GTX113412) and tdTomato fluorescence was used to identify myenteric ganglia. Visualization of the anti-AGO2 and anti-DBT immunoreactivity was performed using anti-rabbit 647 (green) antibodies (1:500) and additional secondary antibodies against human IgG (anti-human 488) was also used for visualizing ANNA1 anti-Hu immunoreactivity. Imaging was performed using confocal microscopy. Tissues were imaged under a 40X oil immersion objective lens, with laser settings selected to ensure no overlap between fluorophores. Images were analyzed using the Fiji ImageJ software. The Johns Hopkins University, and Beth Israel Deaconess Medical Center ACUC approved this work. Johns Hopkins University and the UTHealth IRBs approved these studies, and all individuals provided informed consent.

#### Identification of GI-focused antibodies using murine myenteric plexus

This was performed by immunoprecipitation (IP)/on-bead digest and mass spectrometry as described^5^. LM-MP lysate was used as the antigen source for IP because we were interested in identifying antibodies that target the GI tract. The serum samples were obtained from patients with abnormal GI transit and a distinct immunofluorescence staining pattern of the longitudinal muscle and myenteric plexus (LM-MP) in our cohort of SSc patients with whole gut scintigraphy.

#### Assays to validate autoantibodies and to assay cohorts

Antibodies against AGO and DBT were validated by IP. FLAG-tagged ^35^S-methionine–labeled human proteins generated by in vitro transcription and translation (IVTT) from the appropriate cDNA were used as input for the IPs, which were performed as described previously^5^. In each case, a positive reference IP was performed using an anti-FLAG mouse monoclonal antibody (Sigma). The IVTT-IP assays were subsequently used to screen for these antibodies in banked sera from the cohorts of SSc patients and healthy controls described above.

#### Extracellular vesicle isolation from adult murine LM-MP

Small intestinal LM-MP tissues from a 3-month-old adult male mouse were isolated immediately after the mouse was euthanized and cultured in 5 ml of Stem Cell Media (SCM, containing Neurobasal Medium and B27 supplement; Invitrogen) for 4 hours. The medium was then captured and 0.22 μm filtered (Millipore Sigma). To 5 ml of sterile conditioned SCM, 1 ml of Exoquick-TC solution (Systems Biosciences) was added for EV precipitation. The mixture was gently inverted and refrigerated overnight, then centrifuged at 1500 g for 30 min at room temperature per the manufacturer’s protocol. The supernatant was aspirated, and the pellet was resuspended in 300 μl PBS and stored at −80 °C.

#### Flow analyses of isolated Extracellular Vesicles

The EV suspension was thawed on ice, diluted 1:500 with PBS and analyzed by flow cytometry performed on a Cytoflex nano flow cytometer to assess the particulate size and abundance of EVs.

#### Western blotting

Gel samples made from cultured LM-MP and isolated EVs were electrophoresed on 10% SDS-polyacrylamide gels (50 μg/lane), transferred to PVDF membranes and blotted with an anti-AGO2 antibody (Cell Signaling, Cat. No. 2897, 1:1000 dilution, overnight at 4°C) followed by an anti-rabbit secondary antibody (IR-800 conjugated; LiCor, 1:5000, 2 hours at room temperature). After extensive washing, the blot was imaged using the LiCor Imaging System.

#### Statistical analysis

Clinical and demographic features were compared between patients with and without anti-AGO antibodies using Chi-square or Fischer’s exact tests. Continuous data was compared between groups using Student’s T-tests or Wilcoxon-Mann Whitney according to the type and distribution of the data. Spearman correlation was used to assess the correlation between continuous variables.

STATA 15 (STATA Corporation, College Station, Texas) was utilized for all analyses. A p-value of <0.05 was considered statistically significant.

## RESULTS

### Anti-AGO and anti-DBT antibodies are present in SSc patient sera, but not in healthy controls

Sera from carefully phenotyped SSc patients with GI dysmotility from the GAP cohort were screened by immunofluorescence using LMMP tissue from Wnt1-Cre:tdTomato mice. Sera from 5 patients demonstrated a distinct cytoplasmic punctate staining pattern (representative staining from one of the sera is shown in **Figure 1**) and these were selected for antibody discovery using an IP/on-bead digestion approach^5^. IPs to identify candidate autoantigens were performed using lysate from the adult small intestinal LM- MP of the Wnt1-cre:tdTomato mouse. An IP performed with a healthy control serum was included in the discovery set to identify background hits in the assay run. From the list of potential antigen hits, we prioritized those that were cytoplasmic, that had the most extensive coverage and that were not hits in the control serum IP. We ultimately identified and validated three hits: AGO1 and AGO2 (which we refer to as “AGO”), and DBT. While these autoantigens have been previously reported in patients with SSc^13–15^, recent data suggests that they may have particular significance in the GI tract in patients with dysmotility, given their relevance to extracellular vesicles and mitochondria, respectively^4, 16^. We then screened sera from our SSc all-comers cohort to determine the prevalence of each specificity. 9.4% (17/179) of patients in the all-comers cohort had anti-AGO 1 and/or 2 antibodies (detailed further below), and <1% (1/178) had anti-DBT antibodies. Importantly, none of the healthy controls had anti-DBT antibodies or anti-AGO1/2 antibodies (refer to the Supplemental table). Given the low prevalence of anti-DBT antibodies in the all-comers cohort, further statistical analyses were not performed in this cohort.

**Figure 1.**
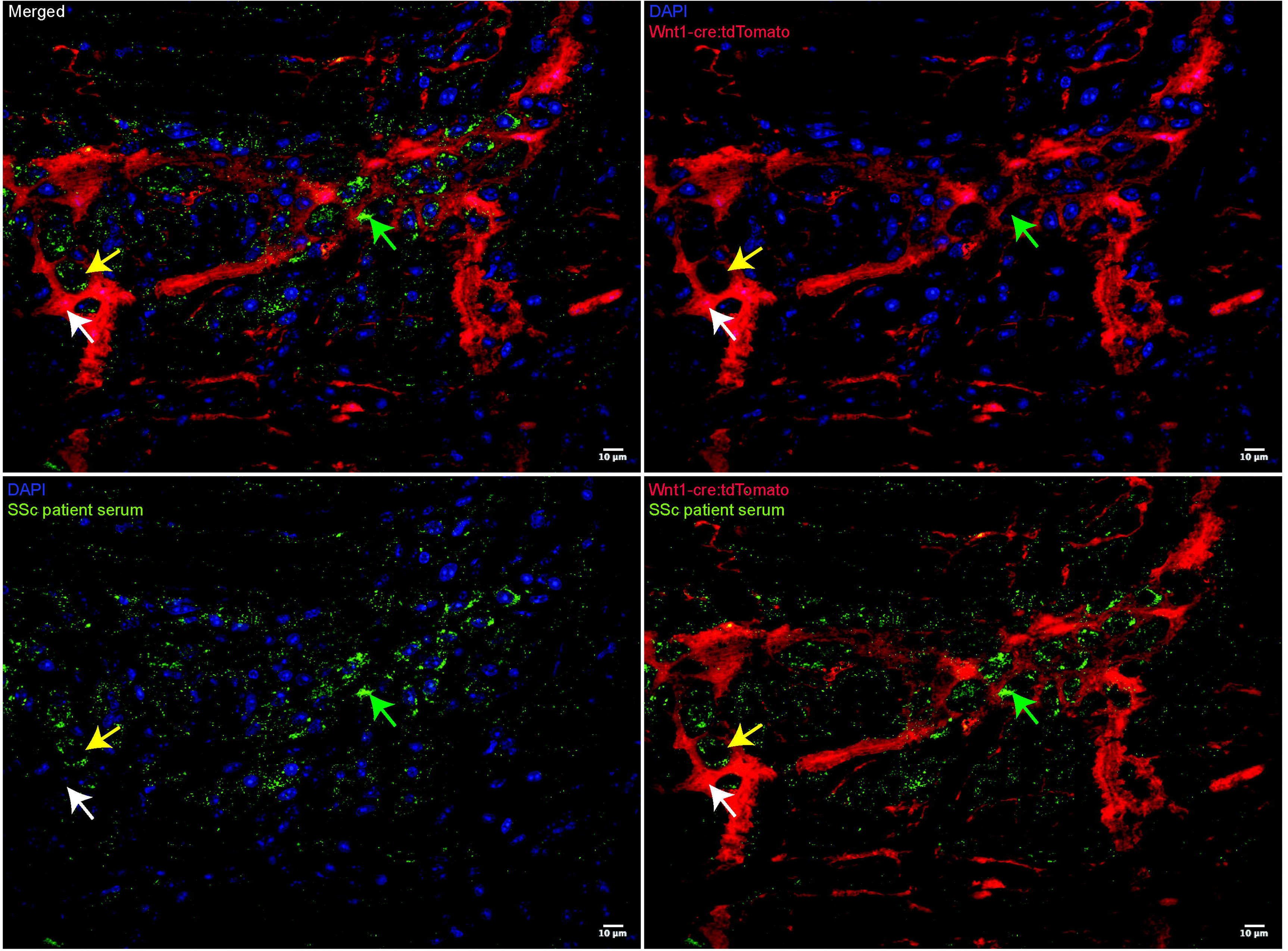
Staining pattern of a representative patient serum on the murine longitudinal muscle and myenteric plexus. Murine ileal myenteric ganglia from an adult Wnt1-cre:tdTomato mouse - where Wnt1-lineage neural crest-derived cells express tdTomato (red) - were immunostained with serum from a patient with SSc and gastroparesis at a dilution of 1:500. Patient autoantibodies (green) target cells within the ganglia that are tdTomato-negative (green arrow), consistent with previously described mesoderm-derived enteric neurons (MENs), as well as occasional tdTomato-expressing neural crest-derived cells (yellow arrow). However, many tdTomato-expressing neural crest-derived cells do not express the autoantigens recognized by SSc serum (white arrow). Autoantibodies in SSc sera also stain cells outside of the myenteric plexus structure. Scale bar = 10 µm.

### Anti-AGO1/2 antibodies associate with GI dysfunction and distress in a SSc all-comers cohort

We then sought to determine whether GI clinical features were associated with anti-AGO antibodies. Of the 17 anti-AGO-positive SSc patients in the all-comers cohort, 17/17 (100%) had anti-AGO1 antibodies, 13/17 (76%) had both anti-AGO1 and anti-AGO2 antibodies, and 0/17 had only anti-AGO2 antibodies. On the UCLA GIT 2.0, the 17 patients with anti-AGO (1 and/or 2) were significantly more likely to have social problems attributable to their GI symptoms, particularly stomach aches and pains [0 (0, 1) vs. 0 (0, 0); p=0.02]. They also trended towards having higher anxiety [1 (0, 2) vs. 0 (0, 1); p=0.05] and fear [0, 1) vs. 0 (0, 0); p=0.06] related to their GI symptoms.

We then looked at the association between GI symptoms and anti-AGO double-positivity in the 13 patients positive for both anti-AGO1 and anti-AGO2 antibodies. We found that the double-positive patients had even more prominent GI symptoms, including significantly more stomachaches/pain [0 (0, 1) vs. 0 (0, 0); p=0.005], socially disruptive bloating [1 (0, 2) vs. 0 (0, 1); p=0.02], and loose stools [1 (0, 2.5) vs. 0 (0, 1); p=0.04] (**Table 1**). They also reported greater emotional distress related to their GI disease, including more worry [1 (0, 2) vs. 0 (0, 1); p=0.012] and fear [0 (0, 1) vs. 0 (0, 0); p=0.02]. Trends were observed for worse overall bloating (p=0.06), more difficulty emptying bowels (p=0.06), and harder stools (p=0.07). No other extraintestinal features were associated with double-positivity in the broader cohort.

**Table 1.** Gastrointestinal symptom severity is associated with anti-AGO1/2 antibodies in the scleroderma all-comers cohort.

|  |  |  |  |
| --- | --- | --- | --- |
| UCLA GIT 2.0 domain and subdomain | **Anti-AGO1/2 positive (n=17)** | **Anti-AGO1/2 negative (n=162)** | **p-value** |
| **Reflux** (total score) | 0.8 (0, 1.9) | 0.4 (0, 0.8) | 0.45 |
| *Acid reflux* | 2 (0, 2) | 0 (0, 1) | 0.07 |
| **Distention** (total score) | 1.3 (0.5, 2) | 0.5 (0, 1.3) | 0.20 |
| *Bloating* | 2 (1, 2) | 1 (0, 1) | 0.06 |
| **Soilage** (total score) | 0 (0, 1) | 0 (0, 0) | 0.43 |
| **Diarrhea** (total score) | 0.5 (0, 1.5) | 0 (0, 0.5) | 0.13 |
| *Loose stools* | 1 (0, 2.5) | 0 (0, 1) | **0.04** |
| **Social functioning** (total score) | 0.3 (0, 1) | 0 (0, 0.3) | 0.08 |
| *Stomach-ache or pain* | 0 (0, 1) | 0 (0, 0) | **<0.01** |
| *Bloated sensation* | 1 (0, 2) | 0 (0, 1) | **0.02** |
| **Emotional well-being** (total score) | 0.3 (0, 0.9) | 0 (0, 0.3) | 0.08 |
| *Feel worried or anxious about your bowel problems* | 1 (0, 2) | 0 (0, 1) | **0.01** |
| *Fear of not finding a bathroom* | 0 (0, 1) | 0 (0, 0) | **0.02** |
| **Constipation** (total score) | 0.8 (0, 1.6) | 0.1 (0, 0.8) | 0.14 |
| *Harder time emptying bowels* | 1 (0, 2.5) | 0 (0, 1) | 0.06 |
| *Hard stools* | 0.5 (0, 2) | 0 (0, 1) | 0.07 |
| **Total GIT score** | 0.8 (0.1, 1.1) | 0.3 (0.1, 0.6) | 0.09 |
| *The Mann-Whitney U test was used to compare the data distribution between groups* |  |  |  |

### Validating the association between anti-AGO antibodies and abnormal GI dysfunction in SSc patients

To validate the association between anti-AGO antibodies and GI dysfunction, we screened a second cohort of SSc patients, which was enriched for significant GI disease (the “GAP cohort”, see Methods section). In this cohort, 13.5% (15/111) had anti-AGO antibodies. Consistent with our findings in the all-comers cohort, all 15 of the antibody-positive patients were anti-AGO1-positive, none had antibodies only against AGO2, and 67% (10/15) had antibodies against both AGO1 and 2. Patients with autoantibodies against the AGO proteins (i.e. either AGO1 alone or AGO1 and AGO2) were significantly more likely to have severe constipation on the UCLA GIT 2.0 [26% vs. 9%; p=0.036; 23% vs. 7%; p=0.035, respectively]. Furthermore, patients who had autoantibodies against both AGO1 and AGO2 were significantly more likely to have severe distention and bloating (16% vs. 4%; p=0.046).

We then performed additional analyses to better understand the relationship between anti-AGO titers and the level of GI severity in SSc patients. Interestingly, we found that patients with severe GI symptoms on the UCLA GIT 2.0 had higher levels of anti-AGO antibodies. Specifically, patients with severe constipation had higher levels of anti-AGO1 and anti-AGO2 antibodies (p=0.03 for both). Furthermore, higher anti-AGO2 antibody levels were observed in patients with severe distention and bloating (p=0.037). Since anti- AGO2 antibodies were not detected in the absence of anti-AGO1 antibodies, these findings suggest that double positivity for anti-AGO antibodies (that is, the presence of both AGO1 and AGO2 antibodies) and higher antibody levels are associated with a severe lower bowel phenotype in SSc.

### Anti-DBT antibodies are associated with slower esophageal transit in SSc patients

In the GI-enriched GAP cohort, we determined that the prevalence of anti-DBT antibodies, while still low, was ∼8 times higher than in the SSc all-comers cohort [4% (4/109) vs. 0.5% (1/178)]. We then explored associations between anti-DBT antibodies and GI symptoms. We found that patients with anti-DBT antibodies had significantly slower esophageal transit at 10 seconds (30% vs. 81%; p = 0.034) and significantly less constipation [0 (0, 0) vs. 0.75 (0.25, 1); p=0.02]. While there were otherwise no significant associations with anti-DBT and symptoms of GI dysfunction, we were likely underpowered to detect more extensive differences.

### Anti-DBT and anti-AGO autoantibodies target cells in and around the Enteric Nervous System

Given that autoantibodies against DBT and AGO co-occur in only a small subset of patients with gastrointestinal dysmotility, and that their target proteins serve distinct cellular functions, we hypothesized that DBT and AGO may be differentially expressed or enriched in distinct cell populations within the enteric nervous system. To investigate this, we examined protein localization in adult murine longitudinal muscle–myenteric plexus (LM–MP) tissue. DBT is a mitochondrial protein and a key component of the branched-chain α-ketoacid dehydrogenase (BCKDH) complex, which plays a critical role in the catabolism of branched-chain amino acids^17^. We recently showed that mitochondria are abundant in a subset of enteric neurons which are derived from the mesoderm, and that autoantibodies against the mitochondrial protein DLAT (also known as the M2 antigen) target the mitochondria-rich MENs in the GI tract. To address whether DBT is similarly enriched in MENs, we immunostained the small intestinal LM-MP tissue with commercial anti-DBT antibodies. We observed that like DLAT/M2, DBT is also enriched in the non-neural crest-derived cells – that is, in MENs (**Figure 2**).

**Figure 2.**
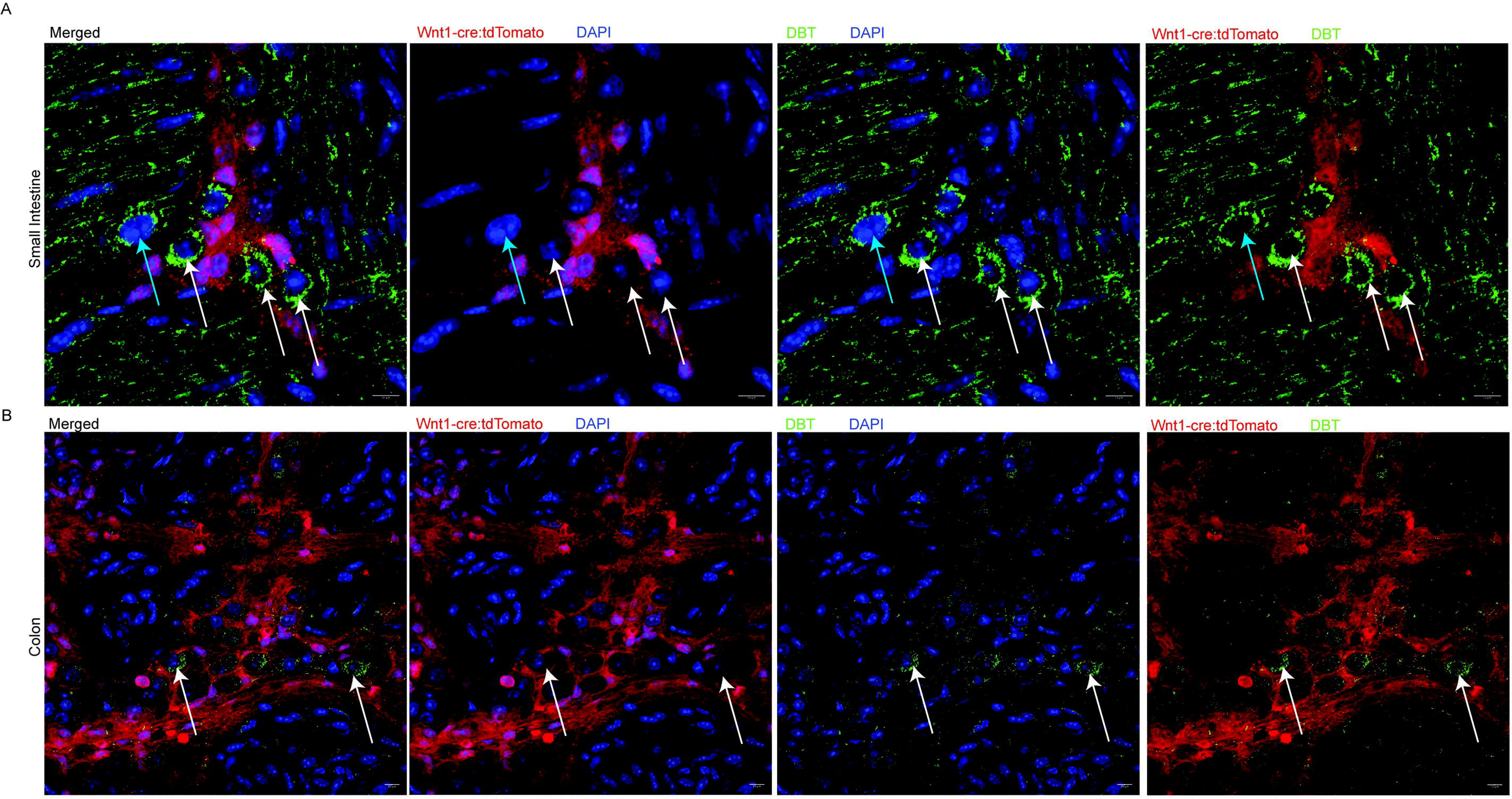
The mitochondrial-specific enzyme DBT is expressed abundantly by mesoderm-derived enteric neurons (MENs) in the adult murine ENS. Representative images of myenteric ganglia from adult murine (A) small intestine and (B) colon show longitudinal muscle–myenteric plexus (LM-MP) tissue isolated from Wnt1-cre:tdTomato mice, in which tdTomato (red) labels all neural crest-derived cells. Tissues were immunostained with commercially available anti-DBT antibodies (green). Within the ganglia, DBT is specifically localized to tdTomato-negative cells (white arrows), previously identified as MENs. Nuclei are stained with DAPI (blue). Scale bar = 10 µm.

AGO1 and AGO2 are essential microRNA (miRNA)-binding proteins, which form a complex with miRNAs to repress protein synthesis by binding to mRNAs and targeting them to RNA processing bodies^18^. AGO proteins and the RNA processing bodies are prevalent in extracellular vesicles (EVs) that are released from cells and have the ability to significantly alter the biology of distant cells, tissues, and organs^19^. EVs have recently been shown to alter GI physiology and are involved in the pathophysiology of GI motility diseases^20^. Both Ago1 and Ago2 are components of the RNA-induced silencing complex (RISC) present in EVs, and Ago2 demonstrates a stronger association with GI symptoms in our patient cohort. Notably, all patients with detectable Ago2 also exhibit Ago1 positivity, whereas the converse is not true. Based on this pattern of expression and its clinical relevance, we elected to immunoblot gut-derived EVs using a specific anti-Ago2 antibody. Therefore, we next investigated whether AGO2 specifically could be detected in EVs released by LM-MP tissues for 4 hours and isolated any released EVs. These were assayed by flow cytometry to test the particle size and abundance. We found that the nominal diameter of EVs was 99.3 nanometers, and that after 4 hours of culture, the adult small intestinal murine LM-MP produced and released 12.4 million/μl EVs (**Figure 3A**). We next tested whether the LM-MP released EVs contained AGO proteins and whether AGO abundance was enriched in the EV fraction compared to the cultured LM-MP tissue.

**Figure 3.**
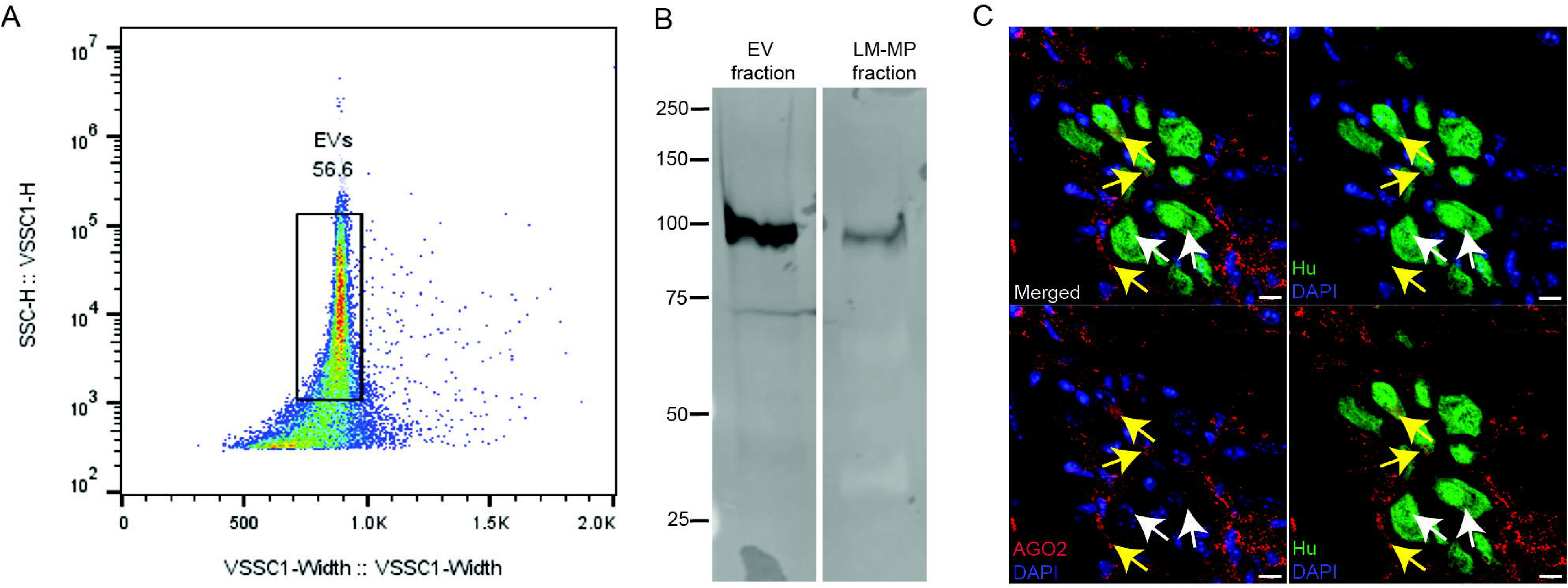
Small intestinal LM-MP tissues release AGO2-enriched extracellular vesicles (EVs), likely of neuronal origin. (A) Short-term cultured adult murine small intestinal LM-MP tissues release abundant EVs into the culture media, as detected by nanoparticle flow cytometry. (B) Western blots of isolated EVs and cultured LM-MP tissue lysates using a commercial anti-AGO2 antibody reveals a strong ∼100 kDa band in the EV fraction and a weaker band in LM-MP lysates, indicating enrichment of AGO2 in EVs. Molecular weight markers (kDa) are shown at left. (C) Immunostaining of adult murine small intestinal LM-MP tissue with neuronal marker Hu (green, white arrows) and anti- AGO2 antibody (red) shows AGO2 signal in and around Hu-positive neurons (yellow arrows), suggesting that enteric neurons are a source of AGO2-containing EVs.

For this, we electrophoresed equal amounts of total protein from the EV fraction and from the LM-MP tissue on SDS-polyacrylamide gels and subsequently immunoblotted using a rabbit monoclonal antibody against AGO2 (**Figure 3B**). A robust 100 kD band was detected in the EV sample while a fainter band was detected in the LM-MP lysate, confirming that AGO2 is present in LM-MP-derived EVs and that it is enriched in the EV fraction. Finally, we tested whether these AGO2-laden EVs were released from the ENS tissues. Since we recently showed that AGO2 is present in and around the cells of the human myenteric plexus^21^, we tested whether AGO2 could be found in and around the cells of the murine ENS. Our data (**Figure 3C**) shows that while the myenteric neurons are moderately immunoreactive against AGO2, AGO2-expressing particles are found on the periphery of the ganglia. This suggests that AGO2-positive EVs may be liberated from the ENS tissue.

## DISCUSSION

Our study demonstrates that antibodies against AGO and DBT associate with distinct GI subtypes in SSc patients. The former are associated with more significant lower GI disease, while the latter are enriched among SSc patients with upper GI disease. Furthermore, the different expression of AGO and DBT in GI and related cells suggests that the mechanisms by which GI dysfunction occurs in these patient subgroups may differ – with the anti-DBT immune response targeting MENs, and the anti-AGO immune response targeting both MENs and EVs produced and released from myenteric ganglia.

We report that autoantibodies against DBT, whose localization follows that of M2/DLAT (another mitochondrial protein in the enteric nervous system^4^), are a newly defined specificity found in SSc patients with GI dysmotility. Consistent with its mitochondrial localization, we observed that DBT is enriched in a specific subset of MENs^4^. DBT protein (the E2 component of the BCKDH complex) catalyzes the first committed step in the oxidative breakdown of the branched-chain amino acids isoleucine, leucine, and valine^17^. The prevalence of anti-DBT in patients with significant GI dysfunction, together with our prior observation that anti-mitochondrial autoantibodies can penetrate viable cells^4^, suggests that anti-DBT autoantibodies may target DBT and lead to mitochondrial dysfunction and GI disease in this subset of patients. While DBT proteins are expressed in multiple other tissues and organs, including in kidneys and heart^17, 22^, the implications of anti-DBT autoantibodies on renal and cardiac functions in SSc patients warrants further investigation.

In contrast, antibodies to AGO proteins may represent a different mechanism of SSc GI disease. Our data show that the LM-MP tissue secretes 10^9^ EVs per ml. The blood plasma concentration of EVs in healthy humans is ∼10^10^ EVs/ml^23^, suggesting that LM-MP tissue accounts for a significant proportion of circulating EVs, which we show express AGO2. AGO proteins are core components of the RNA-induced silencing complex (RISC) and play a central role in RNA interference and microRNA-mediated gene regulation. They mediate gene silencing by binding miRNAs and guiding them to complementary mRNA targets, resulting in translational repression or mRNA degradation^18^. AGO proteins, particularly AGO2 - a key component of the RISC complex - have been identified in EVs where they are thought to facilitate miRNA-based cell-to-cell communication^24^. Anti-AGO2 antibodies may target AGO2-containing EVs, potentially disrupting miRNA-based cell-to-cell communication. Our data shows that the ENS and surrounding cells of the adult LM-MP release AGO2-containing EVs, indicating robust communication within the gut wall and possibly between the gut and other organs. Anti- AGO2 antibodies could therefore interfere with this intra-gut and gut-body signaling network, contributing to disease. Further evidence for the significance of EVs in the maintenance of health and in the development of GI tract diseases is provided in a recent study demonstrating that EVs from the human colon containing long non-coding RNAs play a key role in the development of postinfectious, diarrhea-predominant irritable bowel syndrome^20^. Similarly, a recent review highlights the significance and relevance of non-coding RNA in SSc^25^. While our findings suggest that AGO proteins, and the EVs transporting them, may be targeted by autoantibodies in SSc patients, the mechanisms driving anti-AGO antibody production and the functional consequences of this targeting on the non-coding RNA mechanisms driving SSc await further investigation^17^.

We found that anti-AGO autoantibodies are significantly more associated with psychological complications, including anxiety, fear, and negative social consequences. Interestingly, EV’s arising from gut bacteria have been shown to cross the intestinal barrier and reach the brain, carrying bioactive compounds that influence brain function and emotional health. These vesicles are demonstrated to impact immune responses, neurotransmission, and neurotrophic factors, thereby impacting mood and behavior^26–28^. Furthermore, EVs from the ENS modulate synaptic communication, synaptic strength, and nerve regeneration in the CNS, all of which are critical for maintaining normal cognitive functions and emotional health^29^. These data suggest that the targets of the SSc immune response and the tissues in which they are expressed, may not only provide insight into disease mechanism, but also introduce novel therapeutic targets and lay the foundation for precision-medicine in SSc GI disease.

Our study has several notable strengths. Using gut tissue for autoantibody discovery, we identified DBT as an additional mitochondrial target and determined that gut-secreted EVs containing the AGO-associated RISC complex are a novel target of the autoimmune response in SSc patients. Furthermore, we utilized a large, well-characterized cohort of consecutively recruited SSc patients to define the prevalence and clinical phenotype associated with these specificities, and to determine that antibody levels are associated with disease severity. DBT and AGO expressions were confirmed in murine ENS and surrounding cells, which regulate gut motility. Our study also has some limitations. Since it was cross-sectional, further studies are needed to determine whether these antibodies can be utilized to predict phenotypes in patients with early disease. Future work will also focus on validating these antibodies in other cohorts and determining whether the prevalence of these autoantibodies is related to current or past therapies, and whether targeting AGO proteins alters EV-mediated crosstalk within and between various tissues and organs to cause dysfunction. While our data provides hypothesis-generating associations suggesting a link with disease mechanism, further studies in human models are warranted to confirm a causal link.

## CONCLUSIONS

GI involvement in SSc is clinically diverse and remains insufficiently understood. In this study, we identified one novel autoantibody (anti-DBT) and confirmed the presence of a previously described autoantibody (anti-AGO) in SSc. Importantly, this is the first study to delineate distinct and clinically meaningful GI phenotypes associated with these specific autoantibodies. By mapping the tissue distribution of the corresponding antigens and exploring the potential mechanisms by which these autoantibodies contribute to GI dysfunction, we provide new insights into the possible pathophysiology of SSc-related GI disease. These findings establish a foundation for an antigen-driven framework that may transform our understanding and management of GI manifestations in SSc. Ultimately, this work opens promising avenues for precision diagnostics and targeted therapeutic strategies, paving the way for more personalized care in this complex disease.

## Data Availability

All data produced in the present work are contained in the manuscript

## Acknowledgments and affiliations

We thank Michelle Leatherman, Adrianne Woods and Drs Qingyuan Yang and Laura Gutierrez-Alamillo for their contributions to this work.

---

**Supplemental Table 1.** Characteristics of the SSc patients with and without moderate to high titers of anti-AGO antibodies in the Johns Hopkins SSc Center all-comers cohort

|  |  |  |  |
| --- | --- | --- | --- |
| **Clinical and demographic features** | **Anti-AGO positive**<br>**N=17** | **Anti-AGO negative**<br>**N=162** | **p-value** |
| Age at first symptoms, years, mean (SD) | 57 (11) | 58 (13) | 0.96 |
| Disease duration from first SSc-associated symptom (Raynaud's or non-Raynaud's) to date of serum sample, years, median (IQR) | 10 (5, 17) | 14 (8, 22) | 0.46 |
| Female sex, % (n) | 78 (7) | 87 (148) | 0.35 |
| *Race/Ethnicity* |  |  |  |
| White, % (n) | 89 (8) | 78 (132) | 0.69 |
| Ever smoker, % (n) | 33 (3) | 37 (63) | 1.00 |
| *SSc Type* |  |  |  |
| Diffuse cutaneous disease, % (n) | 33 (3) | 36 (61) | 1.00 |
| Cardiac involvement ($\geq 1$), % (n) | 11 (1) | 16 (27) | 1.00 |
| Myopathy, % (n) | 11 (1) | 24 (40) | 0.69 |
| Sicca symptoms, % (n) | 89 (8) | 78 (131) | 0.69 |
| Raynaud's severity ($\geq 2$), % (n) | 67 (6) | 60 (102) | 1.00 |
| Lung involvement ($\geq 1$), % (n) | 22 (2) | 30 (50) | 1.00 |
| Telangiectasias, % (n) | 100 (9) | 96 (163) | 1.00 |
| Calcinosis, % (n) | 44 (4) | 47 (80) | 1.00 |
| Deceased, % (n) | 11 (1) | 10 (14) | 1.00 |
| *Pulmonary function parameters* |  |  |  |
| FVC, % (n) | 78 (24) | 77 (18) | 0.96 |
| DLCO, % (n) | 53 (35) | 57 (19) | 0.63 |
| RVSP by echo (mmHg), median (IQR) | 36 (35, 45) | 35 (30, 40) | 0.27 |
| *Antibodies, % (n)* |  |  |  |
| Scl-70 (i.e. Topoisomerase-1) | 33 (3) | 21 (36) | 0.41 |
| Centromere (CENP) | 56 (5) | 31 (52) | 0.15 |
| RNA polymerase-3 | 33 (3) | 19 (33) | 0.39 |
| U3RNP | 0 (0) | 10 (17) | 1.00 |
| PMScl | 22 (2) | 12 (21) | 0.33 |
| Ro52 | 22 (2) | 25 (42) | 1.00 |
| ThTo | 0 (0) | 9 (15) | 1.00 |
| Ku | 0 (0) | 7 (11) | 1.00 |
| *SD = standard deviation; IQR = interquartile range; FVC = minimum Forced Vital Capacity & minimum DLCO = Diffusion Capacity on PFT; RVSP = the maximum right ventricular systolic pressure* |  |  |  |

## Supplemental Table

| **Cohort** | **AGO1 (+)** | **AGO2 (+)** | **DBT (+)** |
| --- | --- | --- | --- |
| **All Comers cohort** | 9% (17/179) | 7% (13/179) | <1% (1/178) |
| **GAP Cohort** | 13.5% (15/111) | 9% (10/111) | 4% (4/109) |
| **Healthy Controls** | 0 (0/38) | 0 (0/38) | 0 (0/36) |

## References

1. McMahan ZH, Kulkarni S, Chen J, Chen JZ, Xavier RJ, Pasricha PJ, Khanna D. Systemic sclerosis gastrointestinal dysmotility: risk factors, pathophysiology, diagnosis and management. Nat Rev Rheumatol. 2023;19(3):166–81. Epub 20230206. doi: 10.1038/s41584-022-00900-6. PubMed PMID: 36747090.

2. Furness JB. The enteric nervous system and neurogastroenterology. Nature reviews Gastroenterology & hepatology. 2012;9(5):286–94. Epub 20120306. doi: 10.1038/nrgastro.2012.32. PubMed PMID: 22392290.

3. Kawaguchi Y, Nakamura Y, Matsumoto I, Nishimagi E, Satoh T, Kuwana M, Sumida T, Hara M. Muscarinic-3 acetylcholine receptor autoantibody in patients with systemic sclerosis: contribution to severe gastrointestinal tract dysmotility. Ann Rheum Dis. 2009;68(5):710–4. Epub 20080901. doi: 10.1136/ard.2008.096545. PubMed PMID: 18762475.

4. McMahan ZH, Puttapaka SN, Casciola-Rosen L, Kaniecki T, Gutierrez-Alamillo L, Ming SH, Seika P, Kulkarni S. Antimitochondrial antibodies in systemic sclerosis target enteric neurons and are associated with GI dysmotility. Annals of the Rheumatic Diseases. 2025. doi: 10.1016/j.ard.2025.06.2119.

5. McMahan ZH, Kulkarni S, Andrade F, Perin J, Zhang C, Hooper JE, Wigley FM, Rosen A, Pasricha PJ, Casciola-Rosen L. Anti-Gephyrin Antibodies: A Novel Specificity in Patients With Systemic Sclerosis and Lower Bowel Dysfunction. Arthritis Rheumatol. 2024;76(1):92–9. Epub 20231110. doi: 10.1002/art.42667. PubMed PMID: 37530745; PMCID: PMC10834854.

6. Ayla AY, Kalavar NR, Pimentel M, Morales W, Hummers LK, Shah AA, Hughes M, McMahan ZH. Anti-muscarinic 3 antibodies associate with a severe clinical phenotype in patients with systemic sclerosis. Rheumatology (Oxford). 2025. Epub 20250412. doi: 10.1093/rheumatology/keaf197. PubMed PMID: 40220335.

7. Kulkarni S, Saha M, Slosberg J, Singh A, Nagaraj S, Becker L, Zhang C, Bukowski A, Wang Z, Liu G, Leser JM, Kumar M, Bakhshi S, Anderson MJ, Lewandoski M, Vincent E, Goff LA, Pasricha PJ. Age-associated changes in lineage composition of the enteric nervous system regulate gut health and disease. Elife. 2023;12. Epub 20231218. doi: 10.7554/eLife.88051. PubMed PMID: 38108810.

8. Perin J, Hughes M, Mecoli CA, Paik JJ, Gelber AC, Wigley FM, Hummers LK, Shah AA, Zeger SL, McMahan ZH. Distinct clinical trajectories of gastrointestinal progression among patients with systemic sclerosis. Rheumatology (Oxford). 2025;64(5):2766–74. doi: 10.1093/rheumatology/keae469. PubMed PMID: 39213328; PMCID: PMC12048082.

9. Khanna D, Hays RD, Maranian P, Seibold JR, Impens A, Mayes MD, Clements PJ, Getzug T, Fathi N, Bechtel A, Furst DE. Reliability and validity of the University of California, Los Angeles Scleroderma Clinical Trial Consortium Gastrointestinal Tract Instrument. Arthritis and rheumatism. 2009;61(9):1257–63. doi: 10.1002/art.24730. PubMed PMID: 19714600; PMCID: PMC2767193.

10. Khanna D, Nagaraja V, Gladue H, Chey W, Pimentel M, Frech T. Measuring response in the gastrointestinal tract in systemic sclerosis. Curr Opin Rheumatol. 2013;25(6):700–6. doi: 10.1097/01.bor.0000434668.32150.e5. PubMed PMID: 24047604; PMCID: PMC3880116.

11. Adler B, Hummers LK, Pasricha PJ, McMahan ZH. Gastroparesis in systemic sclerosis: a detailed analysis using whole-gut scintigraphy. Rheumatology (Oxford). 2022;61(11):4503–8. doi: 10.1093/rheumatology/keac074. PubMed PMID: 35136977; PMCID: PMC9629369.

12. Cheah JX, Perin J, Volkmann ER, Hummers LK, Pasricha PJ, Wigley FM, McMahan ZH. Slow Colonic Transit in Systemic Sclerosis: An Objective Assessment of Risk Factors and Clinical Phenotype. Arthritis Care Res (Hoboken). 2023;75(2):289–98. Epub 20220913. doi: 10.1002/acr.24767. PubMed PMID: 34369086; PMCID: PMC8825888.

13. Satoh M, Chan JY, Ceribelli A, Vazquez del-Mercado M, Chan EK. Autoantibodies to Argonaute 2 (Su antigen). Adv Exp Med Biol. 2013;768:45–59. doi: 10.1007/978-1-4614-5107-5_4. PubMed PMID: 23224964.

14. Moadab F, Wang X, Najjar R, Ukadike KC, Hu S, Hulett T, Bengtsson AA, Lood C, Mustelin T. Argonaute, Vault, and Ribosomal Proteins Targeted by Autoantibodies in Systemic Lupus Erythematosus. J Rheumatol. 2023;50(9):1136–44. Epub 20230501. doi: 10.3899/jrheum.2022-1327. PubMed PMID: 37127324; PMCID: PMC10524170.

15. Kuzumi A, Yoshizaki A, Matsuda K, Nagai K, Sato S. Coexistence of systemic sclerosis and cryopyrin-associated periodic syndrome. J Dermatol. 2024;51(9):1240–4. Epub 20240628. doi: 10.1111/1346-8138.17358. PubMed PMID: 38940218; PMCID: PMC11483927.

16. Xu SU, Zhai J, Xu KE, Zuo X, Wu C, Lin T, Zeng LI. M1 macrophages-derived exosomes miR- 34c-5p regulates interstitial cells of Cajal through targeting SCF. J Biosci. 2021;46. PubMed PMID: 34544909.

17. Wang J, Poskitt LE, Gallagher J, Puffenberger EG, Wynn RM, Shishodia G, Chuang DT, Beever J, Hardin DL, Brigatti KW, Baker WC, Gately R, Bertrand S, Rodrigues A, Benatti HR, Taghian T, Hall E, Prestigiacomo R, Liang J, Chen G, Zhou X, Ren L, Liu N, He R, Su Q, Xie J, Jiang Z, Gruntman A, Gray-Edwards H, Gao G, Strauss KA, Wang D. BCKDHA-BCKDHB digenic gene therapy restores metabolic homeostasis in two mouse models and a calf with classic maple syrup urine disease. Sci Transl Med. 2025;17(787):eads0539. Epub 20250226. doi: 10.1126/scitranslmed.ads0539. PubMed PMID: 40009698.

18. Ray S, Roychowdhury S, Chakrabarty Y, Banerjee S, Hobbs A, Chattopadhyay K, Mukherjee K, Bhattacharyya SN. HuR prevents amyloid beta-induced phase separation of miRNA-bound Ago2 to RNA-processing bodies. Structure. 2025. Epub 20250306. doi: 10.1016/j.str.2025.02.003. PubMed PMID: 40056914.

19. Jeppesen DK, Zhang Q, Franklin JL, Coffey RJ. Extracellular vesicles and nanoparticles: emerging complexities. Trends Cell Biol. 2023;33(8):667–81. Epub 20230201. doi: 10.1016/j.tcb.2023.01.002. PubMed PMID: 36737375; PMCID: PMC10363204.

20. Zhou Q, Yang L, Verne ZT, Zhang BB, Fields JZ, Thacker AT, Verne GN. Human colonic EVs induce murine enteric neuroplasticity via the lncRNA GAS5/miR-23/NMDA NR2B axis. JCI Insight. 2025;10(5). doi: 10.1172/jci.insight.178631.

21. Hong SM, Qian X, Deshpande V, Kulkarni S. Optimization of protocols for immunohistochemical assessment of enteric nervous system in formalin fixed human tissue. bioRxiv. 2024:2024.12.15.628584. doi: 10.1101/2024.12.15.628584.

22. Yu JY, Cao N, Rau CD, Lee RP, Yang J, Flach RJR, Petersen L, Zhu C, Pak YL, Miller RA, Liu Y, Wang Y, Li Z, Sun H, Gao C. Cell-autonomous effect of cardiomyocyte branched-chain amino acid catabolism in heart failure in mice. Acta Pharmacol Sin. 2023;44(7):1380–90. Epub 20230329. doi: 10.1038/s41401-023-01076-9. PubMed PMID: 36991098; PMCID: PMC10310802.

23. Johnsen KB, Gudbergsson JM, Andresen TL, Simonsen JB. What is the blood concentration of extracellular vesicles? Implications for the use of extracellular vesicles as blood-borne biomarkers of cancer. Biochim Biophys Acta Rev Cancer. 2019;1871(1):109–16. Epub 20181207. doi: 10.1016/j.bbcan.2018.11.006. PubMed PMID: 30528756.

24. Lv Z, Wei Y, Wang D, Zhang CY, Zen K, Li L. Argonaute 2 in cell-secreted microvesicles guides the function of secreted miRNAs in recipient cells. PLoS One. 2014;9(7):e103599. Epub 20140729. doi: 10.1371/journal.pone.0103599. PubMed PMID: 25072345; PMCID: PMC4114802.

25. Liu Y, Cheng L, Zhan H, Li H, Li X, Huang Y, Li Y. The Roles of Noncoding RNAs in Systemic Sclerosis. Front Immunol. 2022;13:856036. Epub 20220408. doi: 10.3389/fimmu.2022.856036. PubMed PMID: 35464474; PMCID: PMC9024074.

26. Bleibel L, Dziomba S, Waleron KF, Kowalczyk E, Karbownik MS. Deciphering psychobiotics’ mechanism of action: bacterial extracellular vesicles in the spotlight. Front Microbiol. 2023;14:1211447. Epub 20230615. doi: 10.3389/fmicb.2023.1211447. PubMed PMID: 37396391; PMCID: PMC10309211.

27. Guo C, Bai Y, Li P, He K. The emerging roles of microbiota-derived extracellular vesicles in psychiatric disorders. Front Microbiol. 2024;15:1383199. Epub 20240408. doi: 10.3389/fmicb.2024.1383199. PubMed PMID: 38650872; PMCID: PMC11033316.

28. Mottawea W, Yousuf B, Sultan S, Ahmed T, Yeo J, Huttmann N, Li Y, Bouhlel NE, Hassan H, Zhang X, Minic Z, Hammami R. Multi-level analysis of gut microbiome extracellular vesicles-host interaction reveals a connection to gut-brain axis signaling. Microbiol Spectr. 2025;13(2):e0136824. Epub 20241219. doi: 10.1128/spectrum.01368-24. PubMed PMID: 39699251; PMCID: PMC11792502.

29. Zappulli V, Friis KP, Fitzpatrick Z, Maguire CA, Breakefield XO. Extracellular vesicles and intercellular communication within the nervous system. J Clin Invest. 2016;126(4):1198–207. Epub 20160401. doi: 10.1172/JCI81134. PubMed PMID: 27035811; PMCID: PMC4811121.

